# Machine learning prediction of early postpartum prediabetes in women with gestational diabetes mellitus

**DOI:** 10.1101/2023.02.16.23286016

**Authors:** Durga Parkhi, Nishanthi Periyathambi, Yonas Weldeselassie, Vinod Patel, Nithya Sukumar, Rahul Siddharthan, Leelavati Narlikar, Saravanan Ponnusamy

## Abstract

**Background:** Early onset of type 2 diabetes and cardiovascular disease are common complications for women diagnosed with gestational diabetes. About half of the women with gestational diabetes develop postpartum prediabetes within 10 years of the index pregnancy. These women also have double the risk of developing cardiovascular disease than women without a history of gestational diabetes. Currently, there is no accurate way of knowing which women with gestational diabetes are likely to develop postpartum prediabetes. This study aims to predict the risk of postpartum prediabetes in women diagnosed with gestational diabetes.

**Methods:** We build a sparse logistic regression-based machine learning model to learn key variables significant for the prediction of postpartum prediabetes, from antenatal data with maternal anthropometric and biochemical variables as well as neonatal characteristics of 607 UK women diagnosed with gestational diabetes. We evaluate the performance of the proposed model in addition to other more advanced machine learning methods using established metrics such as the area under the receiver operating characteristic curve and specificity for pre-determined values of sensitivity. We use K-L divergence and information graphs to evaluate and compare different thresholds of classification for targeted screening options in resource-constrained settings. We also perform a decision curve analysis to study the net standardized benefit of our model compared to the universal screening approach.

**Results:** Strikingly, our sparse logistic regression approach selects only two variables as relevant but gives an area under the receiver operating characteristic curve of 0.72, outperforming all other methods. It can identify postpartum prediabetes in women with gestational diabetes using the Rule-in test with 92% specificity at an optimal probability threshold of 0.381 and using the Rule-out test with 92% sensitivity at an optimal probability threshold of 0.140.

**Conclusion:** We propose a simple logistic regression model, which needs only the antenatal fasting glucose at OGTT and HbA1c soon after the diagnosis of GDM, to predict, with remarkable accuracy, the probability of postpartum prediabetes in women with gestational diabetes. We envision this to be a practical solution, which coupled with a targeted follow-up of high-risk women, could yield better cardiometabolic outcomes in women with a history of GDM.

## Background

Gestational Diabetes Mellitus (GDM) is defined as any degree of prediabetes with onset or first recognition during pregnancy. Women diagnosed with GDM have upto 10-fold higher risk of Type 2 Diabetes Mellitus (T2DM) compared to those without GDM^1^ and their lifetime risk is around 60% for developing T2DM.^2^ In addition to T2DM, GDM women have a twofold higher risk of Cardiovascular Disease (CVD), at a younger age, and independent of intercurrent T2DM.^3–6^ GDM is associated with an increased risk of cardiovascular dysfunction, including rise in cardiovascular risk factors like blood pressure, and adverse changes in cholesterol and triglycerides.^7^ However, this risk is not the same for all women diagnosed with GDM.

There is some evidence that glucose levels during pregnancy are predictive of prediabetes.^8, 9^ Retnakaran et al.^10^ have shown that the risk of dysglycaemia at 12 weeks postpartum increases across the groups from normal Glucose Challenge Test (GCT) and Normal Glucose Tolerance (NGT), to abnormal GCT and NGT, to Gestational Impaired Glucose Tolerance (GIGT), to GDM. This has been supported by other studies.^11, 12^ Higher fasting glucose shows a high tendency of conversion to T2DM in the postpartum period^7, 13^ and antenatal fasting glucose *>* 5.7 mmol/L is considered to be an important antenatal variable for the prediction of postpartum abnormal glucose metabolism.^14^

Along with glucose values in pregnancy, many studies have proposed the significance of gestational age at the time of diagnosis of GDM, in predicting postpartum prediabetes.^15, 16^ Specifically, women diagnosed at 24 weeks of gestation or earlier, are at higher risk of having postpartum prediabetes.^17^ Similarly, the requirement of insulin therapy during pregnancy, ethnicity, gravidity, BMI, weight at the time of delivery, and neonatal weight, are other factors that have been shown to be associated with the risk of prediabetes.^18^ While there is ample evidence of multiple factors being associated with T2DM onset in GDM-diagnosed women in general, there is no personalized risk score that can predict whether a specific GDM-diagnosed woman is likely to develop prediabetes/T2DM. Indeed, identifying women who are especially at high risk can help in implementing targeted, personalized interventions to delay and prevent the onset of T2DM and its future complications.

Artificial Intelligence has begun to play a dominant role in healthcare, facilitating optimal decision-making as well as personalized treatment. However, its use in the development of predictive models for T2DM onset is still in its nascent stages. Accurate prediabetes risk stratification at or before delivery for GDM women could assist policymakers and clinicians in specifically targeting those at the highest risk, especially in resource-constrained settings. The primary aim of this paper is to investigate the predictive ability of the antenatal variables and derive a model for personalized prediction of prediabetes. We explored the use of logistic regression (LR) and tree-based machine learning algorithms for developing the prognostic model. We report our findings on a multi-ethnic retrospective cohort in the UK. We show that a simple logistic regression model with antenatal fasting glucose and antenatal HbA1c predicts prediabetes in GDM women with a sensitivity of 92% for the Rule-in test and specificity of 92% for the Rule-out test.

## Methods

### Data Acquisition

A retrospective audit of electronic database records of postpartum screening at 6 to 13 weeks of women diagnosed with GDM, from January 2016 to December 2019, was conducted at an NHS trust hospital in the UK. GDM was diagnosed using NICE 2015 criteria.^19^ Complete data is available for 607 women for the following variables: age, height, weight, BMI, systolic and diastolic BP at booking, ethnicity, gravida, parity, smoking status, married status, employment status, gestational age at delivery, mode of delivery, birth weight, breastfeeding status, and biochemical variables such as antenatal fasting glucose (A-FG), antenatal postprandial glucose (A-PG), antenatal HbA1c (A-HbA1c), postpartum fasting glucose (P-FG), postpartum postprandial glucose (P-PG) and postpartum HbA1c (P-HbA1c). Postpartum Oral Glucose Tolerance Test (OGTT) was carried out at 6 weeks and following the change in the NICE guidelines, postpartum HbA1c was carried out at 12-13 weeks following delivery. We define prediabetes as: P-FG ≥ 5.6 mmol/l OR P-PG ≥ 7.8 mmol/l OR P-HbA1c ≥ 40 mmol/mol. ppIFG was defined as P-FG ≥ 5.6 mmol/l and ppIGT was defined as P-PG ≥ 7.8 mmol/l, respectively. We define T2DM as: P-FG ≥ 7.0 mmol/l or P-PG ≥ 11.1 mmol/l or P-HbA1c ≥ 48 mmol/mol.^20^ Normal Glucose Tolerance (NGT) is considered otherwise.

### Statistical Power Analysis

We did a power analysis to determine if the available sample size was sufficient to identify the difference in effect between the normal and prediabetes-diagnosed GDM women. We used the *statsmodels* library and the *TTestIndPower* class in Python to calculate the power analysis for Student’s *t* test for independent samples. For a statistical power of 90%, a minimum sample size of 130 (99 normal and 31 prediabetes) is required for the observed effect size calculated using Cohen’s *d* statistic. We provide the details of power analysis in the Supplementary material.

### Machine Learning

We perform Machine Learning (ML) in Python version 3.7. We compare logistic regression with tree-based methods to build the prognostic model for the prediction of early prediabetes in GDM women. These algorithms inherently address the imbalance in the representation for each of the binary classes of prediabetes outcome using the ‘balanced’ parameter. The ‘balanced’ mode uses the values of *y* to automatically adjust weights inversely proportional to class frequencies in the input data, as the ratio of the total number of samples to the product of the number of classes and the number of occurrences in each class. Mathematically, the class weight is calculated as 1*/*(2 *×* fraction of women in the class). We build the tree-based model using a simple decision tree algorithm, whose performance improves using ensemble methods such as bagging and boosting. All these algorithms use hyperparameters that can significantly affect the performance of these methods on an unseen set. We determine the optimal values of these hyperparameters using nested cross-validation. More specifically, we make the entire data undergo leave-one-out cross-validation (CV1) for model evaluation and we perform an internal stratified 4-fold cross-validation (CV2) on the training folds of CV1 for hyperparameter optimization. We impute the missing values with the Multivariate Imputation by Chained Equations (MICE) technique, using the other non-missing covariates. We scale the training data in CV1 using the *StandardScaler* function and use the *saga* solver in the logistic regression model. The *saga* solver is a variant of the stochastic average gradient (*sag*) solver that also supports the non-smooth L1 penalty, which promotes feature selection. The tree-based algorithms perform feature selection inherently, governed by the optimized hyperparameters in CV2. We perform hyperparameter optimization and model training only on the training folds (*n* − 1 samples) in CV1, with an independent set (1 sample) exclusively held out for testing. We aggregate the model predictions on each held-out sample across the *n* training folds of CV1 and plot the Receiver Operating Characteristic (ROC) curve for this aggregated set. We use the area under the ROC curve as a measure of performance. Finally, we apply it in a similar fashion on the full data to obtain the final model for deployment. We provide the details of the different tree-based methods employed in the supplementary materials.

### Composite Risk Score calculation

Using the coefficients from the final fitted logistic regression model on the full data, we develop a composite risk scoring system using the best selected antenatal variables to predict the probability of prediabetes in GDM-diagnosed women. We calculate the composite risk score as the probability of class 1 obtained from the logistic regression model. It is given by the expression 1*/*(1 + *e*^−*b*^), where *b* = *b*_0_ + *b*_1_ *·x*_1_ + *b*_2_ *·x*_2_ + … + *b*_*m*_ *·x*_*m*_ where *b*_0_ is the intercept and *b*_*m*_ coefficient of mth variable (*x*_*m*_), respectively.

We compute specificity, positive predictive value (PPV), negative predictive value (NPV), accuracy, and the F1 score at five predetermined values of sensitivity (60%, 70%, 75%, 80%, and 90%) for the optimal selected model. We give the definition/formulae for all these in the supplementary section.

### Kullback-Leibler (K-L) Divergence and Information Graphs to evaluate and compare diagnostic tests and select optimal cut-point

We use the information theory approach in Lee et al.,^21^ Samawi et al.,^22^ and Benish et al.,^23^ briefly summarized below, to select the optimal probability threshold for accurate prediction of the binary outcome of prediabetes. An important approach followed in medical diagnostics is to predict the ‘Rule-in and Rule-out’ potential of the diagnostic test to safely include the patients in need of treatment and discard those not in need, respectively. At a probability threshold *c* reported by the ML algorithm, suppose the proportion of the diseased population correctly predicted as diseased is given by *g*_1_(*c*) and that of the non-diseased population correctly predicted as non-diseased is given by *g*_2_(*c*). Both *g*_1_(*c*) and *g*_2_(*c*) are Bernoulli probability distributions and are simply the sensitivity and specificity, respectively at the threshold value of *c*. The K-L divergence (or relative entropy) measures the separation between these two probability distributions and is given by:

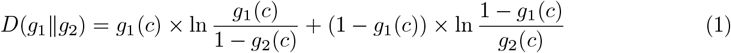

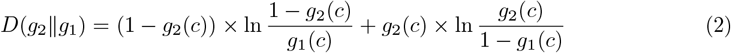

By definition, *D*(*g*_1_‖*g*_2_) ≥ 0, *D*(*g*_2_‖*g*_1_) ≥ 0. The KL divergence is close to 0 when there is little difference between the two distributions. A high *D*(*g*_1_‖*g*_2_) value indicates the increase in information of predicting disease onset. We calculate *D*(*g*_1_‖*g*_2_) and *D*(*g*_2_‖*g*_1_) for 1000 cut points at an interval of 0.001 from 0 to 1. We chose *T*_in_ with cut-point *c*_in_ corresponding to *D*_max_(*g*_1_‖*g*_2_) as the diagnostic test with greatest rule-in potential. We chose *T*_out_ with cut-point *c*_out_ corresponding to *D*_max_(*g*_2_‖*g*_1_) as the diagnostic test with greatest rule-out potential. We calculate 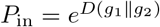, which is the ratio of post-test odds to the pre-test odds of having the disease for a randomly selected diseased individual. We also calculate 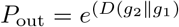, which is the ratio of pre-test odds to the post-test odds of having the disease for a randomly selected non-diseased individual. *P*_in_, *P*_out_ ≥ 1.

Next, we calculate the Information Distinguishability measure, 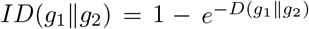 and 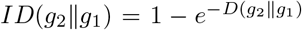, to study and compare the separation provided by the diagnostic test between the diseased and the non-diseased distributions. We calculate the objective function *TKL*_discrete_(*c*) = *D*(*g*_1_‖*g*_2_) + *D*(*g*_2_‖*g*_1_) and chose the optimal cut-point *c*_in−out_ corresponding to max(*TKL*_discrete_(*c*)) to achieve maximum information for *T*_in−out_ with high potential in both rule-in and rule-out situations. Further, we plot information graphs to characterize and compare the performance of our diagnostic tests at different cut-points depending upon the rule-in or rule-out potential. The expected value of the relative entropy provides a measure of the expected diagnostic information and plotting it as a function of the pre-test probabilities yields an information graph. The equations used to plot the information graphs are given as follows: Let *D*_i_ be the true status and *T*_i_ be the diagnostic test result for the patient, respectively, (*i* = *{*0, 1*}*, 0: disease absent, & 1: disease present). If *x* = *Pr*(*D*_1_), then the diagnostic information obtained from a +ve, and -ve test result (*I*_+_(*x*), *I*_−_(*x*), respectively) and the expected diagnostic information (IE(x)) are given as follows.

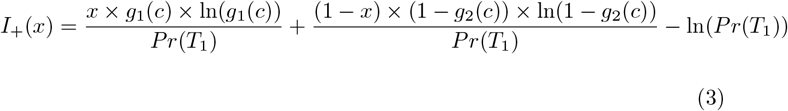

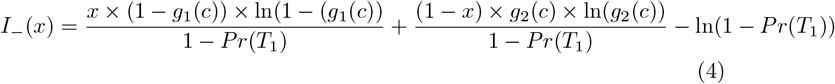

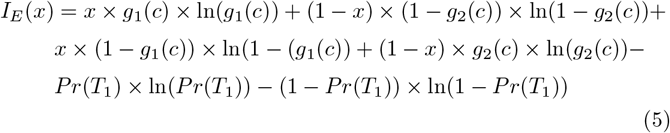

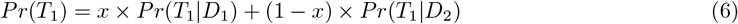

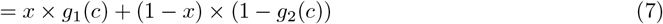

In addition, we also plot the information graph by representing the total K-L divergence as the discrete Bregman divergence, which is the sum of the vertical distances between the negative Shannon entropy function (see Supplementary material for details) and tangents to it at probabilities *p* = *g*_1_(*c*) and *p* = 1 − *g*_2_(*c*).

### Decision Curve Analysis

We carry out Decision Curve Analysis (DCA) to evaluate and compare the performance of our model in comparison to the ‘treat all’ and ‘treat none’ approaches. Finally, we compare the correctly identified non-attenders (sensitivity) vs follow-ups avoided (the true negatives + false negatives, obtained from the optimal selected model), to calculate the number of women requiring enhanced care, to maximize targeted postpartum follow-up.

## Results

Postpartum glucose status was available for 394 (64.91%) out of the 607 women. 340 (56.01%) women underwent OGTT at 6 weeks and 128 (21.09%) underwent the postpartum HbA1c around 13 weeks. prediabetes is present in 92 (23.35%) women. Of these 47 (51.09%) were abnormal by P-FG, 33 (35.87%) by P-PG, and 39 (42.39%) by P-HbA1c. We show the baseline characteristics of these 394 women in Table 1.

**Table 1:** Comparison of antenatal, delivery and postnatal characteristics of GDM women with presence and absence of prediabetes

| Variable | All<br>(N=394) | prediabetes<br>(N=92) | ppNGT<br>(N=302) |
| --- | --- | --- | --- |
| **Maternal characteristics** |  |  |  |
| Age | 32.21 ± 5.40 | 32.38 ± 5.46 | 32.16 ± 5.39 |
| Height (m) | 1.64 ± 0.07 | 1.64 ± 0.07 | 1.64 ± 0.07 |
| Weight (kg) | 79.78 ± 19.80 | 84.32 ± 22.82 | 78.36 ± 18.58 |
| BMI ($kg/m^2$) | 29.76 ± 6.81 | 31.21 ± 7.40 | 29.30 ± 6.56 |
| Systolic BP (mmHg) | 115.71 ± 13.62 | 116.07 ± 13.78 | 115.60 ± 13.59 |
| Diastolic BP (mmHg) | 69.98 ± 9.40 | 70.41 ± 8.18 | 69.85 ± 9.76 |
| **Parity** |  |  |  |
| 1 | 192 (48.98%) | 43 (46.74%) | 149 (49.67%) |
| >= 2 | 200 (51.02%) | 49 (53.26%) | 151 (50.33%) |
| **Ethnicity** |  |  |  |
| White European | 303 (76.90%) | 66 (71.74%) | 237 (78.48%) |
| South Asian | 46 (11.68%) | 13 (14.13%) | 33 (10.93%) |
| Others | 45 (11.42%) | 13 (14.13%) | 32 (10.60%) |
| **Smoking category** |  |  |  |
| Never smoked | 190 (50.94%) | 43 (49.43%) | 147 (51.40%) |
| Ex-smoker | 147 (39.41%) | 34 (39.08%) | 113 (39.51%) |
| Smoker | 36 (9.65%) | 10 (11.49%) | 26 (9.09%) |
| **Marrital Status** |  |  |  |
| Single | 21 (5.74%) | 3 (3.45%) | 18 (6.45%) |
| **Employment own/partner** |  |  |  |
| Unemployed | 9 (2.56%) | 3 (3.53%) | 6 (2.25%) |
| **At OGTT and Intrapartum** |  |  |  |
| GA at antenatal OGTT (weeks) | 28.16 ± 4.21 | 27.50 ± 4.08 | 28.37 ± 4.23 |
| A-FG (mmol/L) | 4.95 ± 0.87 | 5.38 ± 0.91 | 4.82 ± 0.81 |
| A-PG (mmol/L) | 8.55 ± 1.75 | 8.90 ± 1.75 | 8.44 ± 1.74 |
| A-HbA1c (mmol/mol) | 35.52 ± 4.69 | 38.13 ± 4.61 | 34.72 ± 4.42 |
| GA birth (weeks) | 37.91 ± 1.27 | 37.65 ± 1.28 | 37.99 ± 1.26 |
| Preterm (GA ≤ 37 weeks) | 53 (13.59%) | 18 (20.00%) | 35 (11.67%) |

| Table 1 – continued |  |  |  |
| --- | --- | --- | --- |
| Variable | All<br>(N=394) | prediabetes<br>(N=92) | ppNGT<br>(N=302) |
| **Delivery mode** |  |  |  |
| Spontaneous | 197 (50.38%) | 37 (40.66%) | 160 (53.33%) |
| Instrument assisted | 32 (8.18%) | 6 (6.59%) | 26 (8.67%) |
| Caesarean delivery | 162 (41.43%) | 48 (52.75%) | 114 (38.00%) |
| **Neonatal characteristics** |  |  |  |
| Birth weight (grams) | 3211.95 ± 467.75 | 3216.48 ± 511.41 | 3210.57 ± 454.58 |
| **Birth Centile** |  |  |  |
| AGA (10-90th centile) | 267 (74.58%) | 61 (72.62%) | 206 (75.18%) |
| SGA (< 10 centile) | 42 (11.73%) | 10 (11.90%) | 32 (11.68%) |
| LGA (> 90 centile) | 49 (13.69%) | 13 (15.48%) | 36 (13.14%) |
| Male baby | 183 (46.80%) | 42 (46.15%) | 141 (47.00%) |
| Breastfeeding initiated | 207 (58.31%) | 45 (54.88%) | 162 (59.34%) |
| **Postpartum maternal biochemical characteristics** |  |  |  |
| P-FG (mmol/L) | 4.99 ± 0.62 | 5.64 ± 0.79 | 4.78 ± 0.38 |
| P-PG (mmol/L) | 5.59 ± 1.62 | 7.10 ± 2.08 | 5.12 ± 1.07 |
| P-HbA1c (mmol/mol) | 37.53 ± 4.84 | 42.22 ± 4.56 | 34.99 ± 2.55 |

### Machine Learning Analysis

The data is imbalanced (as expected), with a 23.35% representation of the positive prediabetes class. We compare simple logistic regression with different classification tree methods for predicting prediabetes from training on this small and imbalanced data set. We use class-weight = balanced in the logistic regression algorithm and ‘balanced’ classification tree-based algorithms from the imbalanced-learn python package for developing the tree-based prognostic models. The predictive performance of our proposed framework improves significantly by applying ensemble methods of bagging and boosting to the base decision tree estimator but remains lower than LR. LR gives the area under the ROC curve of 0.7203 from aggregating the test predictions from the leave-one-out cross-validation (Fig 2a). Using the base decision tree algorithm and leave-one-out cross-validation, the area under the ROC curve for the aggregated test predictions is 0.6210, bagging decision trees improves it to 0.6883. Random forests further improve it to 0.6944 using 4-fold stratified cross-validation in CV1 and the maximum area under the ROC curve from the tree-based algorithms is 0.6991 from balanced bagging using histogram-based gradient boosting tree classification algorithm using 4-fold stratified cross-validation. We use 4-fold stratified cross-validation in CV1 instead of leave-one-out for random forests and the boosting algorithm due to the high time complexity of leave-one-out. We conclude that the simplest prediction algorithm for binary classification, LR, outperforms the advanced tree-based methods in the prediction of prediabetes. Our final composite risk score using the LR model with A-FG and A-HbA1c is highly robust for the prediction of prediabetes in GDM women. Out of the n=394 runs of leave-one-out cross-validation, antenatal fasting glucose and antenatal HbA1c are selected 318 (*>* 80%) times.

**Figure 1:**
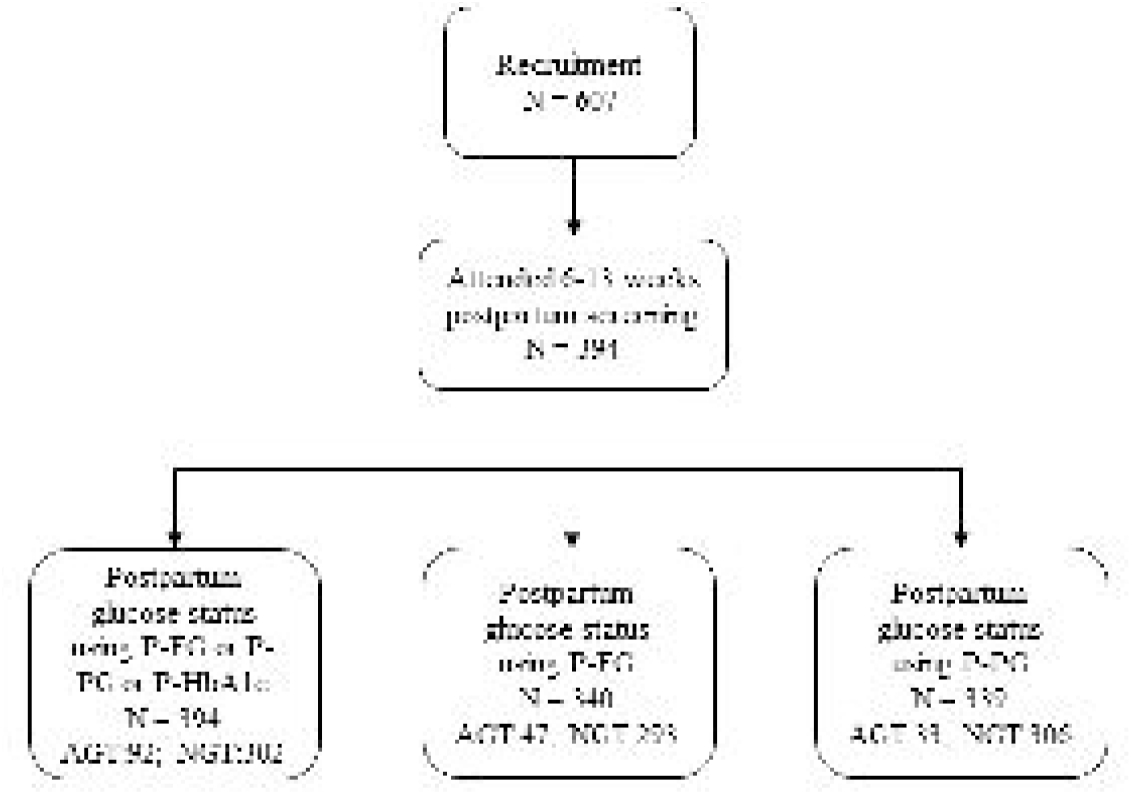
Consort diagram of early postpartum glucose tolerance. Caption: The flow chart displays the proportion of GDM women with and without prediabetes. The diagnosis of prediabetes was made if: FPG ≥ 5.6 or 2-hr glucose ≥ 7.8 at postpartum OGTT or HbA1c ≥ 40 mmol/mol.

**Figure 2:**
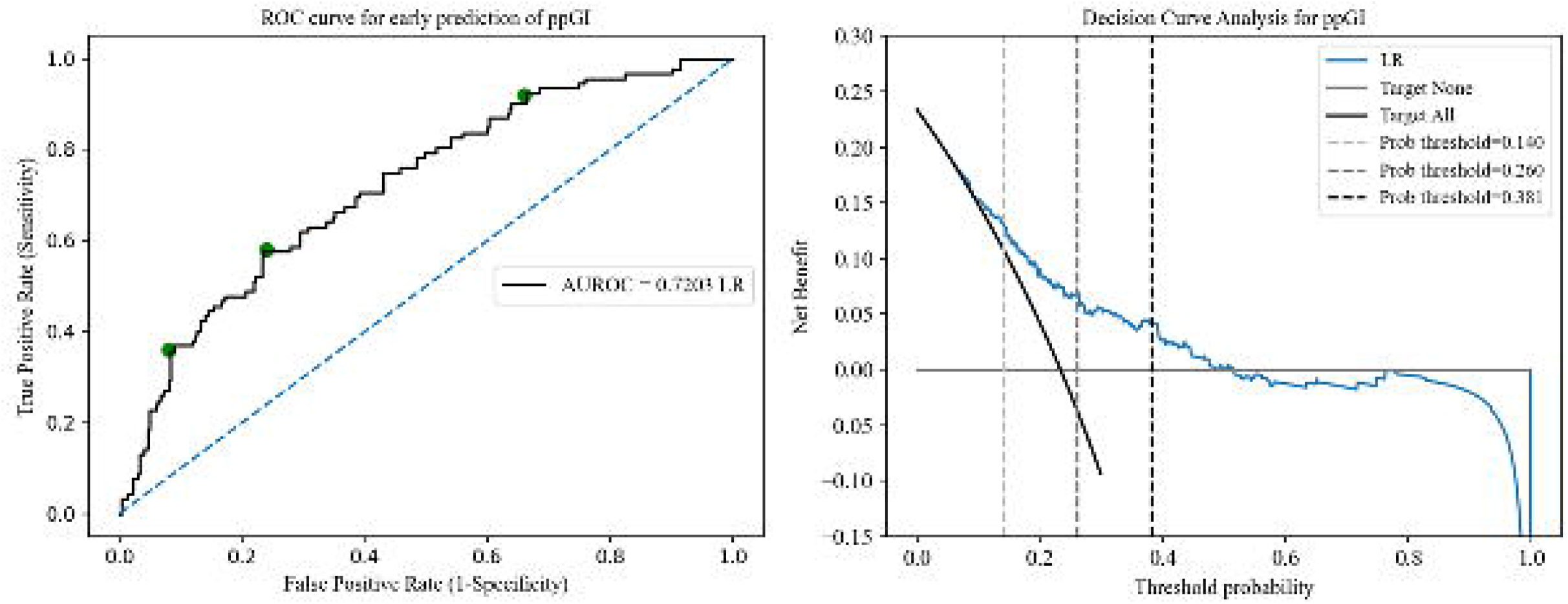
A. Estimated ROC for the prediction of postpartum prediabetes following a GDM pregnancy using ML model B. Decision curve analysis for the standardized net benefit obtained for the optimal probability threshold. Caption: Fig (a): AUROC (Area under the receiver operating characteristic) was used to evaluate the performance of our machine learning-based method using the logistic regression model on the validation cohort, n= 394 by aggregating the predictions from the test folds of CV1. The area under ROC was 0.7203. The green dots on the ROC curve represent *T*_in_ (*c*_in_ = 0.381), *T*_in−out_ (*c*_in−out_ = 0.260), and *T*_out_ (*c*_out_ = 0.140), from left to right, respectively. Fig (b): The DCA (Decision curve analysis) showed the net benefit obtained from the ML (blue) prediction model. The net benefit of implementing our model in a clinical setting is larger when compared to the follow-up of all GDM women for prediabetes. DCA was derived from the equation, Net benefit 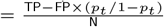, where TP and FP are the true positives and false positives respectively, *p*_*t*_ is the probability threshold, and N is the total number of participants in the validation cohort, n=607.

### Composite Risk Score calculation

Based on our proposed final Logistic regression model, we calculate the composite risk score, *c* (or P(prediabetes)), as

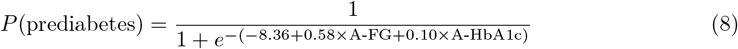

### Kullback-Leibler (K-L) Divergence and Information Graphs to evaluate and compare diagnostic tests and select optimal cut-point

*T*_in_ with *D*_max_(*g*_1_(*c*), *g*_2_(*c*)) = 0.30 and *c*_in_ = 0.381 has high specificity of 92%, in concurrence with the ‘Rule-in-specific-test’ principle and *T*_out_ with *D*_max_(*g*_2_(*c*), *g*_1_(*c*)) = 0.28 and *c*_out_ = 0.140 has high sensitivity of 92%, again in concurrence with the ‘Rule-out-sensitive-test’ principle. *P*_in_ = 1.35 and *P*_out_ = 1.23 for *T*_in_, and *P*_in_ = 1.21 and *P*_out_ = 1.33 for *T*_out_, which is the increase (decrease) in disease odds after the test for a diseased (control) individual. *T*_in−out_ with max(*TKL*_discrete_(*c*)) = 0.51 for *c*_in−out_ = 0.260 has *P*_in_ = 1.31 and *P*_out_ = 1.27. Also, maximum of the Youden’s index, *J*_max_ = 0.34 (*J* (*c*) = *g*_1_(*c*)+*g*_2_(*c*)−1), and maximum *F*_1_-score = 0.49 occurs at the same *c*_in−out_ = 0.260. 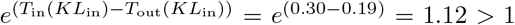, which implies that positive result obtained by *T*_in_ is more likely to be true than positive result obtained by *T*_out_. In other words, *T*_in_ is more specific and yields fewer false positives compared to *T*_out_. Similarly, 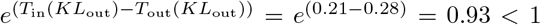 shows that *T*_in_ is less sensitive with more false negatives.

We generated the information graphs using the equations for *I*_+_(x), *I*_−_(x), and *I*_E_(x) as a function of *x* = *Pr*(*D*_1_), as shown in Fig 3a, 3b, and 3c. We can observe that *T*_in_ provides the most diagnostic information when the test result is positive, and the pre-test probability of a positive result (*Pr*(*D*_1_)) is low. *T*_out_ provides the most diagnostic information when the test result is negative, and the pre-test probability of a positive result is high. For *T*_in−out_, we obtain more diagnostic information when the test yields a positive result than a negative one and we obtain maximum information from a positive result at a lower pre-test probability than that from the negative result. In Fig 3d, we can see the information gained using the discrete Bregman divergence representation of *TKL*_discrete_ by adding the vertical distances from the negative Shannon Entropy function to the tangents drawn at probability *p* = *g*_1_(*c*) and 1 − *g*_2_(*c*).

**Figure 3:**
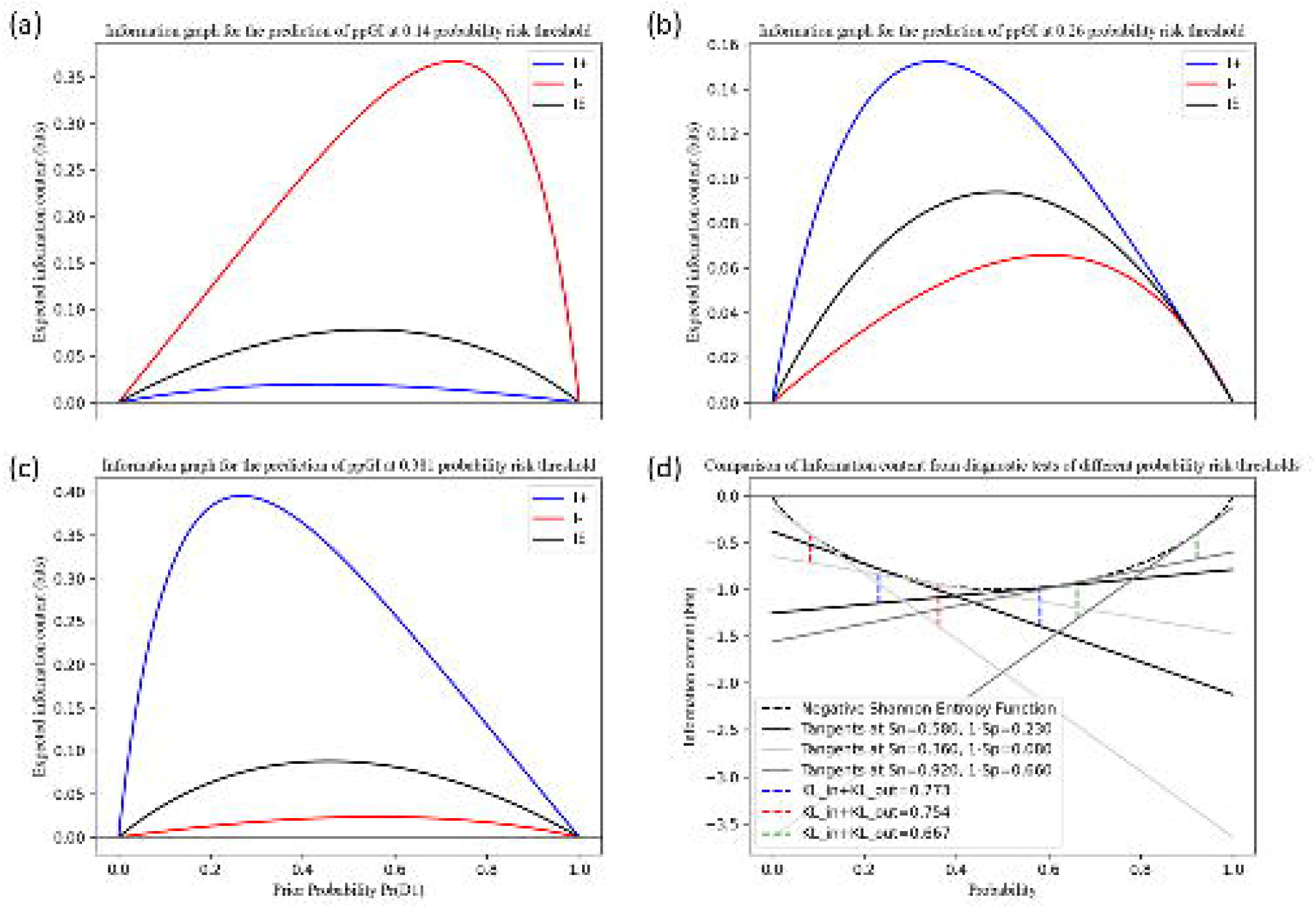
Information graphs for comparing Rule-in and Rule-out test potentials of diagnostic tests using different cut-points for predicting a low and high risk of prediabetes in GDM women Caption: Information graphs provide means to distinguish between diagnostic test performance. We compared the diagnostic information obtained from *T*_out_, *T*_in−out_, and *T*_in_ defined by the cut-points 0.140, 0.260, 0.381. A positive diagnosis made by the ‘Rule-in-specific-test’ and a negative diagnosis made by the ‘Rule-out-sensitive-test’ gives us the most information, as expected. Fig (a), (b), (c): Maximum information from a positive test diagnosis (blue) is obtained at a lower pre-test probability than the maximum information from a negative test diagnosis (red). The diagnostic test with a lower cut-point gives maximum information when the diagnosis is negative (i.e, the test is very sensitive and we can rule out the negative cases safely) and the diagnostic test with a higher cut-point gives maximum information when the diagnosis is positive (i.e, the test is very specific to the disease and we can rule in the positive cases safely). *I*_E_ is the expected information from the diagnostic test (*x × I*_+_ + (1 − *x*) *× I*_−_, where *x* is the probability of a positive test diagnosis). Fig (d): The sum of the distances between the tangents to the negative Shannon entropy function at *p* = *g*_1_ (*c*) and *p* = 1 − *g*_2_ (*c*) is the discrete Bregman divergence, which represents total K-L divergence.

Using the prognostic model with LR, 15 out of 100 women are above the optimal threshold of 0.381, and focusing on these women could improve the early prediabetes diagnosis. 28 out of 100 women are below the optimal threshold of 0.140, and testing for early prediabetes diagnosis can be safely avoided in this category. The model shows 92% sensitivity for the rule-in test and 92% specificity for the rule-out test, Table 3 shows the sensitivity, specificity, PPV, NPV, F1 score, accuracy, and other measures related to K-L divergence at different probability thresholds.

**Table 2:** Factors associated with postpartum prediabetes by machine learning model

| variables | $\beta$ (SE) | OR (95% CI) | p-value |
| --- | --- | --- | --- |
| A-FG (mmol/L) | 0.5816 (0.207) | (0.175, 0.988) | 0.005 |
| A-HbA1c (mmol/mol) | 0.0996 (0.038) | (0.025, 0.174) | 0.009 |

**Table 3:**
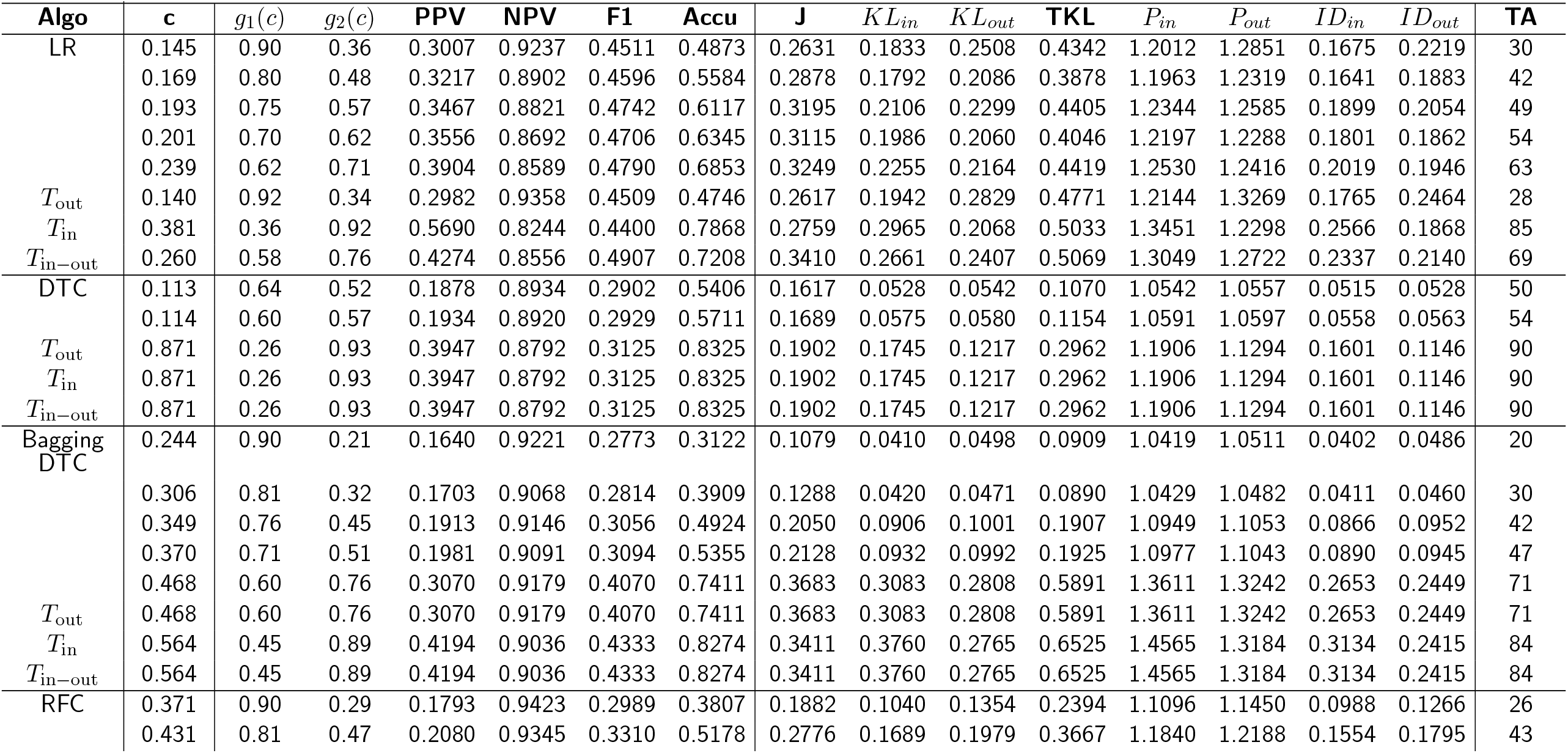
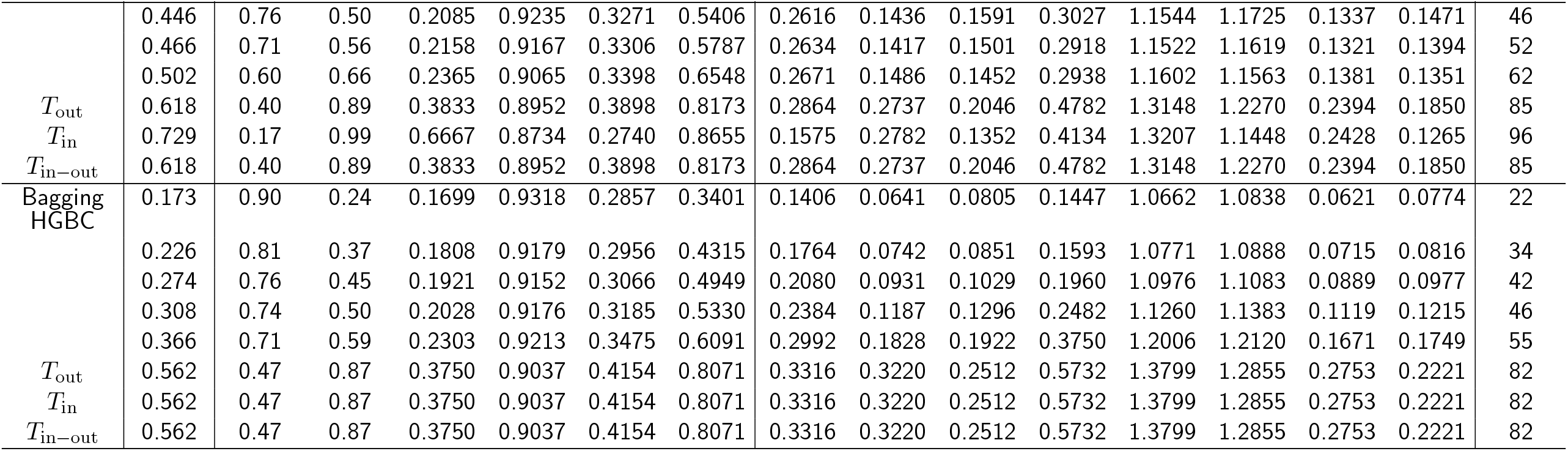
Performance of the diagnostic test for postpartum prediabetes at various probability thresholds

### Decision Curve Analysis

In the decision curve analysis by comparing the ‘treat all’ and ‘treat none’ approaches, the ML model obtains a higher standardized net benefit as compared to the universal screening of all GDM women for early prediabetes (Fig 2b).

## Discussion

In this study, we try to predict at the time of delivery if the women diagnosed with GDM are at high risk of getting diagnosed with postpartum prediabetes at 6-13 weeks postpartum. For this purpose, we employ a variety of machine learning techniques including both logistic regression and advanced tree-based algorithms and train the models using routinely collected antenatal and delivery variables as predictors. Our proposed model using nested cross-validation and logistic regression algorithm can effectively predict prediabetes in GDM women, using only the antenatal predictors fasting glucose and HbA1c, with good sensitivity and specificity.

The use of machine learning for predicting postpartum prediabetes in GDM-diagnosed women has been rarely studied. We are aware of only one study that has made use of machine learning algorithms to predict the occurrence of T2DM post-GDM: Krishnan et al.^24^ proposed random forest and gaussian naive Bayes algorithms to predict T2DM after GDM, and achieved a modest specificity of 23% at a sensitivity of 88%. It also lacked the use of advanced techniques to deal with imbalanced data. Real-world medical data is scarce due to the different challenges posed in its collection. To the best of our knowledge, there is no larger data collected for studying prediabetes in GDM women than the data in the present study. In our study, we propose a more personalized approach to identifying postpartum prediabetes after GDM, at the antenatal visit itself, by calculating a simple score based on only two easy-to-measure biochemical predictors, obtained using machine learning techniques and a logistic regression algorithm, with good sensitivity and specificity (each of 92% for Rule-out and Rule-in tests, respectively). Further, we suggest different cut-offs for classifying high-risk women depending upon resource availability.

The proposed prediction test needs only the antenatal fasting glucose (at the time of antenatal OGTT) and HbA1c, usually measured soon after the diagnosis of GDM for clinical use. Thus, no additional tests/costs are involved, and is easy to use by healthcare professionals. The information theory analysis proposes different cut-offs for classification according to the requirement of ruling-in or ruling-out the prediabetes condition in GDM-diagnosed women. All women diagnosed with GDM during pregnancy are recommended to have annual screening,^19, 25^ although the compliance is currently poor.^5, 26^ Therefore, we can allow for more false positives than false negatives and propose *c*_out_ = 0.140 as the optimal cut-off for classification. However, in low-resource settings, we can primarily focus on women with *P* (prediabetes) ≥ *c*_in_ = 0.381 and then consider women with *P* (prediabetes) ≥ *c*_in−out_ = 0.260 in the following step. If resource constraint is not an issue, we can target women with *P* (prediabetes) ≥ *c*_out_ = 0.140 as well. Targeting GDM women stepwise according to their risk of developing prediabetes is more personalized than the blanket approach of targeting all women with GDM. This could be a pragmatic approach in settings with limited resources. The desired cut-off out of *c*_in_, *c*_out_, or *c*_in−out_ can be chosen depending upon the purpose and setting in which this diagnostic test is used.

Postpartum weight loss has been shown to reduce the risk of incidence of T2DM and recurrent GDM in the subsequent pregnancy.^27, 28^ However, initiating such lifestyle interventions can be difficult due to lack of personalization and may not produce optimum results due to poor adherence by the women.^29^ Our approach to identifying women with a high risk of prediabetes (using any ‘*c*’) can provide an improved understanding of individualized prediabetes risk which can be used to target women for interventions (diet and lifestyle, encourage breastfeeding, etc) for postpartum weight loss. This can in turn improve their T2DM and CVD risk profile. Women are most conducive to interventions during pregnancy and also maintain close contact with healthcare professionals. Identifying the high-risk women during the antenatal visits will help the healthcare professionals to implement necessary interventions throughout the remaining pregnancy period, and also encourage postpartum follow-up.

We believe that the results obtained are supportive for testing and validating our Rule-in and Rule-out composite risk score approach on a larger prospective dataset.

The key strength of our study is the use of a variety of machine learning techniques and the comparison of the LR algorithm with tree-based algorithms for developing the prognostic model for individualized risk prediction of prediabetes following GDM pregnancy. In addition, to the best of our knowledge, this is the first study that used K-L divergence and information graphs for evaluating and comparing different diagnostic tests at different cut-points and explaining their rule-in and rule-out potentials. However, our study has important limitations. First, this is retrospective data and hence other potential variables that could influence the prediabetes status such as gestational weight gain and insulin treatment were not electronically available. Second, postpartum glucose status data was only available in 65.0% of the cohort, although this follow-up rate for postpartum glucose testing was higher than the national average. Finally, while the sample size is small (*n* = 394 and *n* = 92 for the prediabetes class) for machine learning analyses, this was adequate based on the substantial predictive performance and the power calculations. In addition, the only available literature to our knowledge that looked at predicting the onset of T2DM following GDM was based on only 77 patient records with 15 variables.^24^

## Conclusions

This study shows that our proposed model using a logistic regression algorithm is effective for the prediction of prediabetes in GDM women by using the already available antenatal fasting glucose and antenatal HbA1c. We believe that this approach is easy for practical use with no additional cost and could be extremely effective for individualized risk stratification of GDM women. This approach could be used for targeted glucose testing during the postpartum period in a resource-constrained setting.

## Data Availability

The full dataset is available upon request following the completion of a suitable confidentiality agreement.

## Declarations

## Acknowledgements

This study was conducted as an audit at the George Eliot Hospital NHS Trust, Nuneaton, UK. We extend our thanks to Mrs. J Wilson, Mrs. J Plester, Mrs. T Ritchie, and Mr. S Selvamoni for their help in providing the list of all the GDM women. Mrs. T Ritchie is the advanced nurse practitioner who facilitates the postpartum testing.

## Funding

PS and YW are partly funded by Medical Research Council, UK (MR/R020981/1); DP is funded by Warwick-Novo Nordisk international Doctoral Training Program and NP’s Ph.D. is funded by Chancellor’s International Scholarship, University of Warwick. RS is supported by his institution, funded by the Department of Atomic Energy, Government of India.

## Abbreviations

GDM: Gestational Diabetes Mellitus
T2DM: Type 2 Diabetes Mellitus
CVD: Cardiovascular Disease
OGTT: Oral Glucose Tolerance Test
LR: Logistic Regression
ROC: Receiver Operating Characteristic
DCA: Decision Curve Analysis

## Math variables used in the paper

n: Sample size (n = 607)

x: variable

m: Number of variables (m = 25)

y: class label

## Ethics approval and consent to participate

This study was carried out as an audit and quality improvement project using an anonymous dataset. The local clinical governance team approved the audit and formal ethical approval was not required.

## Competing interests

The authors declare that they have no competing interests.

## Consent for publication

Not applicable.

## Authors’ contributions

1. Conceptualization: DP, LN, RS, and PS
2. Data curation: DP, NP, NS, YW
3. Formal analysis: DP
4. Funding acquisition: PS
5. Investigation: NP, PS
6. Methodology: DP, PS, LN, RS
7. Project administration: VP, NS, PS
8. Resources: VP, NS, PS
9. Software: DP
10. Supervision: PS, LN, RS
11. Writing - original draft: DP
12. Writing - review & editing: All authors

## Notes

### Competing Interest Statement

The authors have declared no competing interest.

