## Supplemental Table 1 for "Machine learning prediction of early postpartum prediabetes in women with gestational diabetes mellitus"

**Supplementary Material**

**SA. Power Analysis**

In statistics, power analysis is used to determine the probability of finding a significant difference between two sample distributions, if it exists. A statistical hypothesis test makes an assumption about the outcome. The null hypothesis in a statistical test is that there is no significant difference between specified populations, any observed difference is due to sampling or experimental error. The statistical power is the probability of correctly rejecting the null hypothesis. Therefore, in mathematical terms, power can be defined as probability of True positives (TP). For a predefined significance level and known effect size, we can either fix power and calculate minimum required sample size to obtain the desired effect or calculate power for the available sample size.

Antenatal fasting (ANF) and antenatal HbA1c (ANHbA1c) are the two selected predictors for antenatal prediction of prediabetes in GDM diagnosed women. The sample distributions for ANF and ANHbA1c for the GDM (class 1) and non-GDM (class 0) groups are as shown in Figure S1(a) and S1(b), respectively. Let r be the ratio of the number of samples in the second sample distribution to those in the first. Then r = Nobs2/Nobs1 = 92/302 = 0.305.

Fig S1: (a) The sample distributions for ANF (b) The sample distributions for ANHbA1c

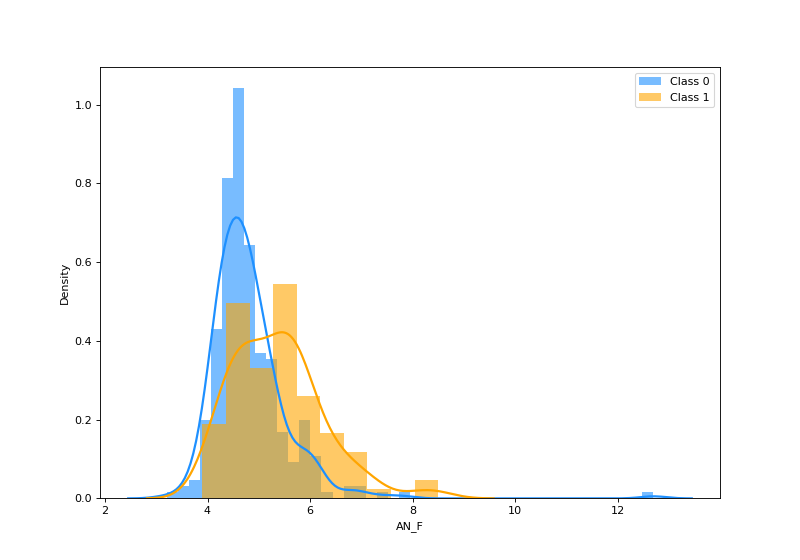

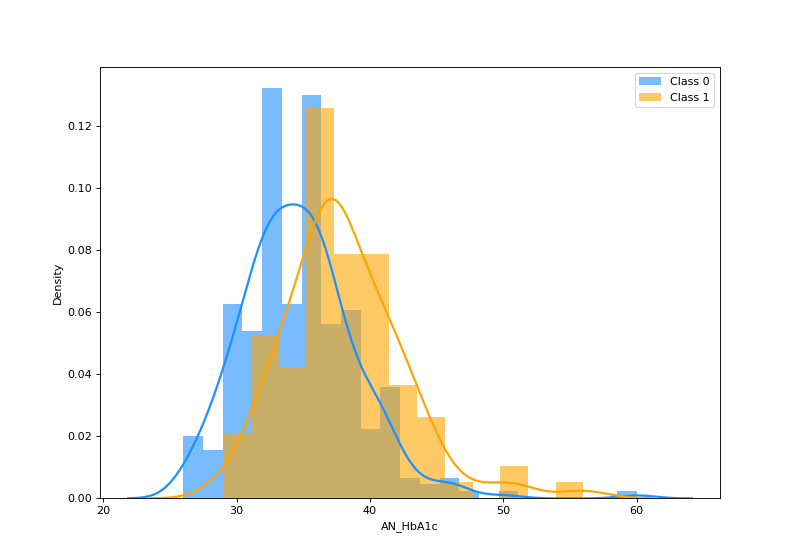

Calculating effect size

We will use the Cohen's *d* for calculating the effect size. Let 𝑛₁, and 𝑛₂ be the number of samples in distribution 1 (class 0) and distribution 2 (class 1), respectively. Let μ₁ and μ_2_ be the means and $\sigma$₁ and $\sigma$_2_ be the standard deviations of the two sample distributions. Then, the Cohen's d statistic is given by (Cohen, Jacob. Statistical power analysis for the behavioral sciences. Academic press, 2013.):

d = $\frac{\mu_{1}-\mu_{2}}{\sqrt{\frac{\left( n_{1}-1 \right).\sigma_{1}^{2}+\left( n_{2}-1 \right).\sigma_{2}^{2}}{n₁+n₂-2}}}$.

Assuming the sample distributions of ANF and ANHbA1c for class 0 and class 1 are normal, we get d_ANF_ = 0.681 and d_HbA1c_ = 0.781.

Calculating Sample size for fixed Power

Let us fix significance level α=0.05 and statistical power *p*=0.9. Using the Cohen’s d calculated above, we get the minimum required sample size as 130 (99 class 0 + 31 class 1) for ANF and 99 (76 class 0 + 23 class 1) for ANHbA1c.

Lastly, we plotted power curves to see how the power of the test changes with the other parameters: sample size, effect size, and significance level. In Figure S2, we can see how the power of the test increases with increasing sample size, for different fixed effect sizes. We can understand that if the effect size is small (greater overlap between the two sample distributions), then greater number of observations are required to identify the existing significant difference between the two sample distributions, and thus correctly reject the null hypothesis. Also, the power of the test increases with increasing effect size.

Fig S2: Change in power of the test as a function of sample size, for fixed effect sizes

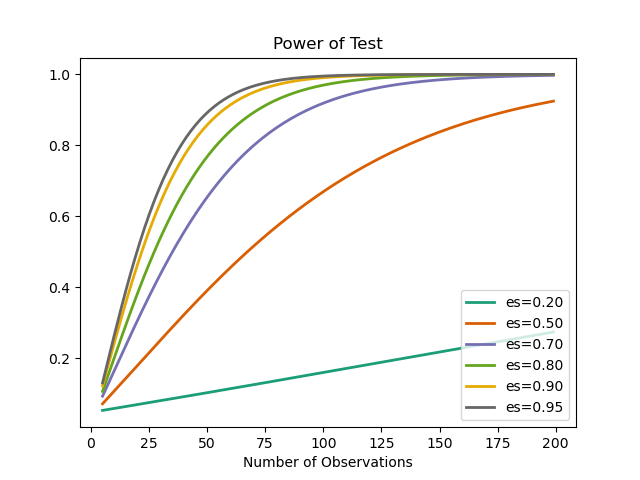

**SB. Concept Diagram of the Machine Learning model**

Fig S3: Concept diagram of our proposed method. Replace leave-one-out with 4-fold stratified cross validation for the model using Random forests and boosting algorithms.

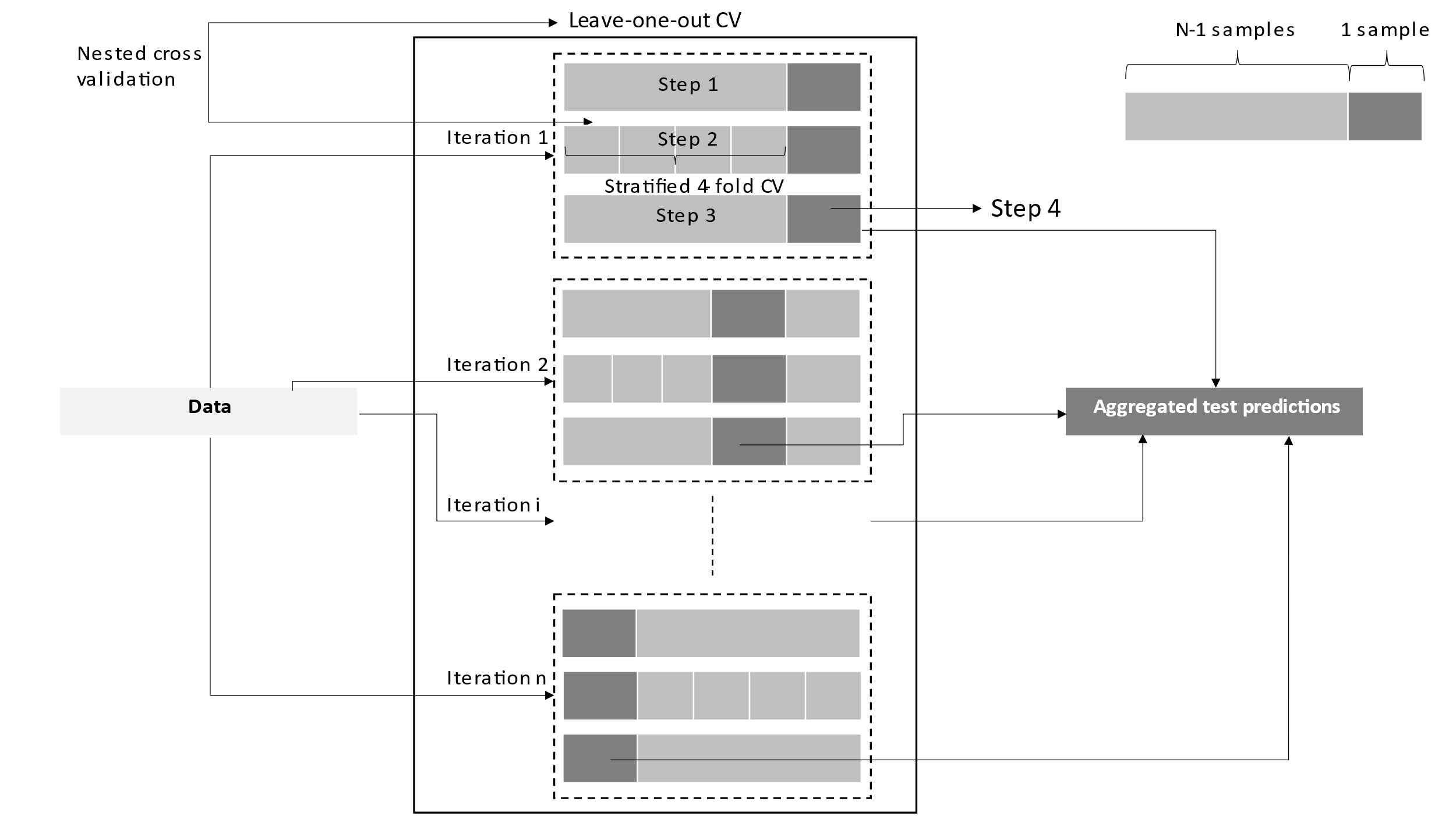

**SC. Details of the tree-based algorithms**

***Balanced Decision Tree***

A single decision rule is developed from learning from the training data in each iteration i of CV1 and is used to make predictions on the held-out test data. The number of features used in the decision rule, their order, the splitting cut-offs at each node in the decision tree, etc. are decided by optimizing the hyperparameters: [*max_leaf_nodes, min_samples_split, min_samples_leaf, criterion*] in CV2.

***Balanced Bagging using Decision Tree***

When the training data is small, *b* different bootstrapped training data sets can be generated by sampling with replacement from the original training data. The model is trained on each of these *b* training data sets to get f^b^(x) and the final classification model is obtained by averaging all the *b* predictions, f_bag_(x) = $\frac{1}{B}\sum_{b=1}^{B} f^{b}\left( x \right)$. f_bag_(x) is used to make predictions on the held-out test data. The decision tree hyperparameters optimized are same as above.

***Balanced Random Forest***

Random forests are similar to bagged decision trees except for the number of features considered at each split in the decision tree- all features are split candidates in bagged decision trees vs a random sample of *m* predictors are the split candidates in random forests. The hyperparameters for optimization are similar as for decision trees, except that criterion is replaced by *m* which is either *sqrt* or *log2*.

***Balanced Bagging using Histogram-based Gradient Boosting Tree***

Boosting works in a similar fashion to bagging, however the individual decision trees are grown sequentially using information from previously grown trees, and on modified version of the original training data set. The hyperparameters optimized in this method in CV2 are: [*max_leaf_nodes, min_samples_leaf, max_depth, l2_regularization*].

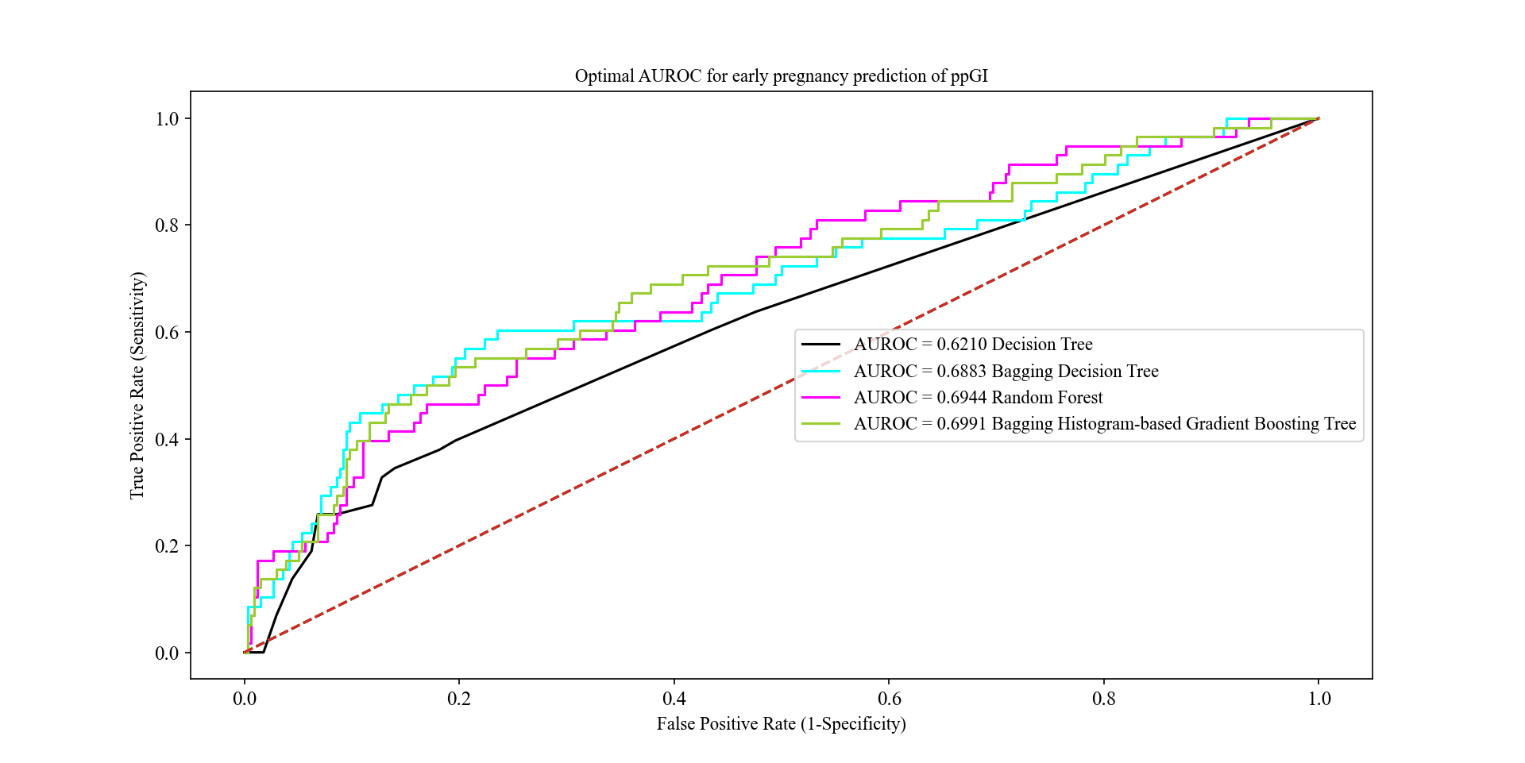
Fig S4: ROC curve for early prediabetes prediction using tree-based methods

Fig S5: Variation in the mean stratified 4-fold cross validation accuracy as a function of the lasso regularization hyperparameter C for the final model. Maximum CV-accuracy of 0. 8554 is obtained for C=0.0798

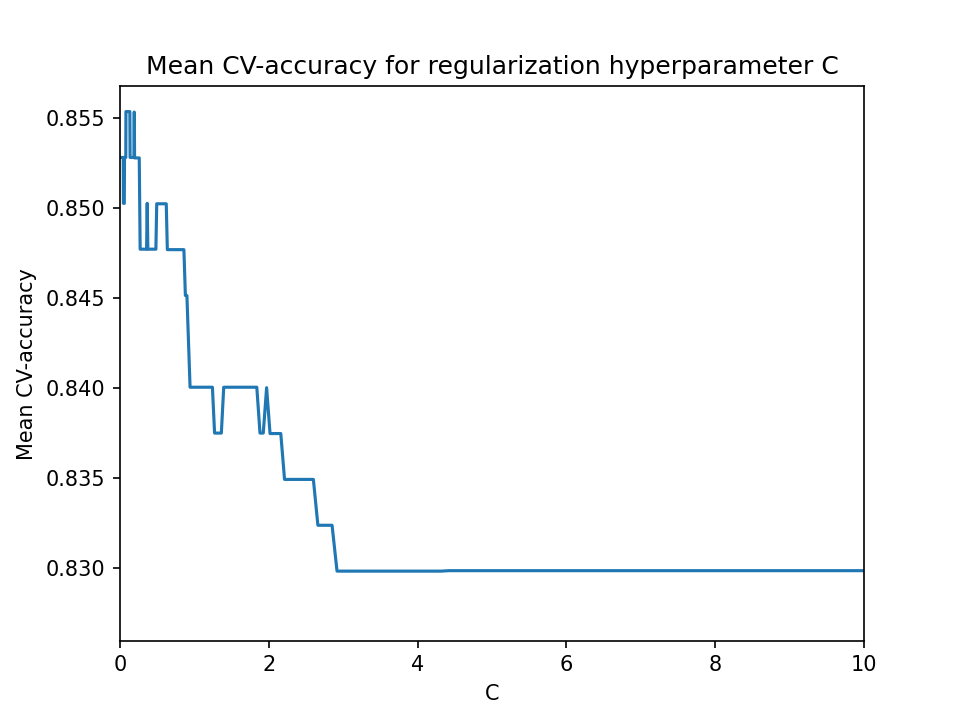

The LR model could predict ppIFG with an area under the ROC curve of 0.6598.

Fig S6: ROC curve for early ppIFG prediction using LR

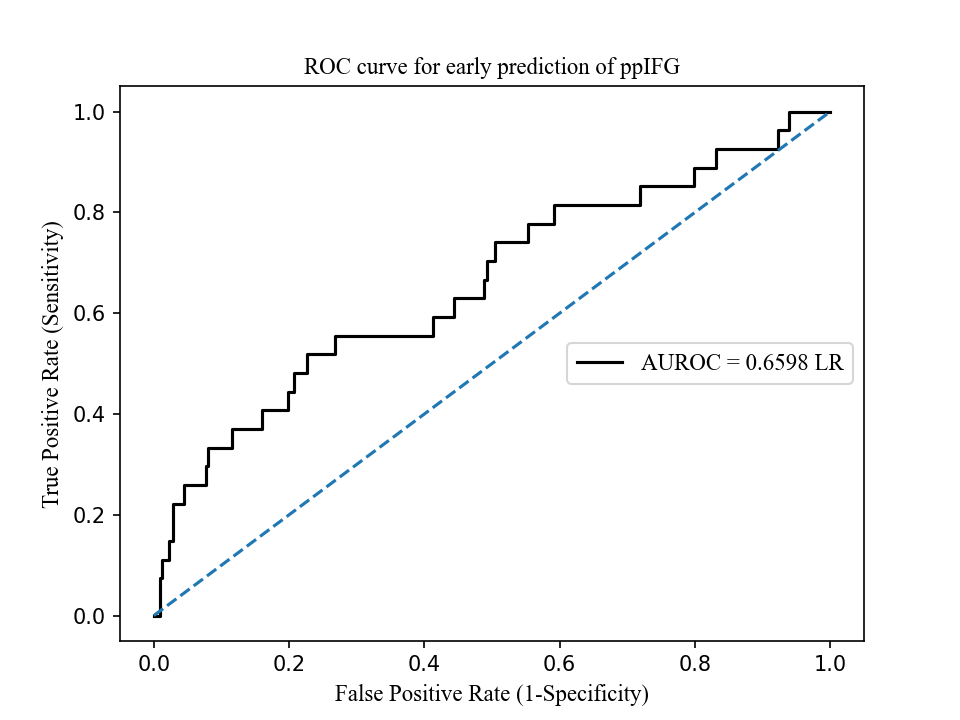

**SD. Basic formulae**

F1 score: 2 × Precision × Recall/(Precision + Recall)

Negative Shannon entropy function: h(p) = p × ln (p) + (1-p) × ln (1-p)

Appendix 1: Standard letter format for postpartum screening to all GDM women issued from the Diabetes clinic, GEH-NHS Trust, UK.

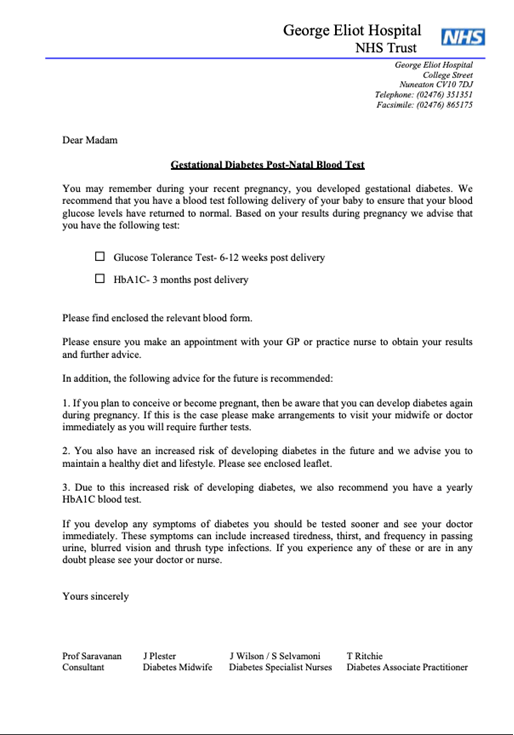
